# COVID-19 vaccination, from first dose to booster: New insights into the frequency of most common systemic adverse events and possible booster nocebo effects based on a systematic review

**DOI:** 10.1101/2021.12.15.21267847

**Authors:** Claudia Behrens, Maria Samii-Moghadam, Tatiana Gasperazzo, Anna M. Gross, Jack Mitchell, Johannes B. Lampe

**Affiliations:** Lampe & Company GmbH & Co. KG, Berlin, Germany

**Author notes:** Corresponding author: Johannes B. Lampe, Lampe & Company GmbH & Co. KG, 10553 Berlin, Germany.

## Abstract

**Background:** Based on placebo data, it has been recently demonstrated that the frequencies of most common adverse events (AEs) of COVID-19 vaccination are overestimated due to negative expectation bias of vaccine recipients (nocebo effect). Since booster studies lack comparators, estimating the extent of the nocebo effect is difficult. We aimed to overcome this obstacle through a systematic comparison of most common AE frequencies across vaccine doses (first, second, booster), age groups, and vaccine vs. placebo arms.

**Methods:** We systematically assessed systemic AEs in approved COVID-19 vaccines according to the PRISMA guidelines. All documents regarding COVID-19 vaccines with a booster dose authorized by the FDA (cutoff date 19 November 2021) were systematically searched on PubMed and the FDA website. Solicited systemic AEs from all documents supporting approval/authorization were collected. After standardization of doses and age groups, AE frequencies were compared between vaccine and placebo.

**Findings:** Two trials were identified for BNT162b2 (n=21,785 participants), two for mRNA-1273 (n=22,324), and one for Ad26.COV2.S (n=4,085). Fever cases dropped to about half with the booster dose in all vaccines, whereas all other systemic AE frequencies were similar to the preceding dose. Almost no fever cases occurred with placebo (first/second dose); all other systemic AEs occurred at high frequencies. After subtracting placebo arm values from vaccine values, the frequencies for the various AEs were roughly comparable within each dose for each vaccine.

**Interpretation:** Fever is the only solicited systemic AE that can be assessed objectively. It occurs about 50% less often with the booster than with the preceding dose. This may indirectly indicate a considerable overestimation of systemic AEs in the case of booster vaccinations and a pronounced nocebo effect. The nocebo effect appears to substantially contribute to the differences in the frequencies of the various systemic AEs.

**Funding:** No funding received.

**Research in context:** *Evidence before this study:* A high nocebo effect has recently been shown for the most common solicited adverse events (AEs) based on the randomized controlled trials of several vaccines approved for COVID-19. To date, there has been no systematic review of systemic AEs in COVID-19 vaccines with a booster dose approved by the FDA. Normally, assessing the extent of the nocebo effect requires the implementation of randomized, placebo-controlled studies. However, for ethical reasons, studies underlying the authorization of COVID-19 booster vaccines are not controlled. Therefore, reported AEs lack any reference parameter. We searched PubMed and medRxiv for nocebo effects regarding the objectively measurable AE fever using the terms “(COVID) AND (nocebo) AND (fever)” with no language restrictions; no hits were found on PubMed and six were found on medRxiv. The search without any indication, using only the search terms “(nocebo) AND (fever)”, resulted in two hits on PubMed and six hits on medRxiv. An additional search was performed for “(vaccine or COVID) AND nocebo”; 21 hits were found on PubMed and 12 on medRxiv. None of the hits obtained with the three searches showed evidence of relevance for the present topic of a direct association of fever with the nocebo effect.

*Added value of this study:* Instead of placebo AE data as reference parameter, we suggest fever as an objectively measurable value to estimate the extent of the nocebo effect in uncontrolled vaccination studies. To our knowledge, this is a new concept with which evidence for the extent of a possible nocebo effect in COVID-19 booster vaccination can be provided. With this approach we directly supplement information on the most recently reported nocebo effects in first, second and booster vaccine doses. To the best of our knowledge, this is the first study to indirectly compare the solicited systemic AE frequencies of approved COVID-19 vaccines across all doses, with easy-to-understand graphs and based on all approval-relevant trials. This is astonishing, as in the context of a pandemic it is essential to communicate scientific data in a generally understandable manner and according to the highest scientific standards. The graphs provided in this work could serve as an example for such clear communication.

*Implications of all the available evidence:* Using the example of systemic AEs, we were able to show that the frequency of AEs in COVID-19 booster vaccination may be overestimated. Fever as an objective measure to estimate the nocebo effect could help to optimize public awareness campaigns in the future. The graphic presentation of results facilitates a deeper understanding of complex scientific data and provides new insights. It is therefore ideal for a highly dynamic scientific field and may also be applied to other challenges in the COVID-19 pandemic and beyond.

## Introduction

Vaccination is an important tool to combat viral infections. In the case of COVID-19, the approved vaccines are well tolerated and most adverse events (AEs) are only mild^1^. Even more, the results of a recent study suggest that a substantial proportion of the AEs following COVID-19 vaccination may be due to a negative expectation bias of participants, the so-called nocebo effect^2^ assessed in placebo arms. This study was based on the initial approval randomized controlled trials (RCTs), which are the best sources of efficacy and safety data, as they provide control data as reference points. Therefore, these studies are essential to better estimate the relevance of more frequent AEs.

In this sense, the evaluation of AE frequencies after booster COVID-19 vaccines with a current Emergency Use Authorization (EUA) in the USA (BNT162b2, mRNA-1273 and Ad26.COV2.S) is particularly challenging, since the booster studies are not controlled and the reported AEs thus lack any reference parameter.

We have recently published the results of the first approach that enables an indirect comparison of approved COVID-19 vaccines using all available RCT data^3^. Here we apply the same approach to AEs occurring after the booster dose. This may help to objectify the current discussions around the safety of booster vaccines against COVID-19 and reduce the hesitancy among the public towards receiving a booster dose. Moreover, it serves as an outstanding example of how an indirect comparison among different clinical studies can be accomplished by means of systematic, standardized assessments of the biomedical literature that goes beyond traditional reviews.

## Material and Methods

### Study selection

The PRISMA 2020 Statement^4^ was adopted for study selection and data collection. A systematic search on PubMed and FDA website was performed. Three vaccines with a booster dose have an EUA in the USA: the mRNA vaccines BNT162b2 (Comirnaty) by Biontech/Pfizer and mRNA-1273 (Spikevax) by Moderna, and the adenovirus vector vaccine Ad26.COV2.S by Janssen. All journal articles and regulatory documents which formed the basis for the EUA of the booster dose were identified. All publications available as of 19 November 2021 reporting results of these trials were considered and included in the assessment. Study identification was performed independently by two authors, according to the predefined eligibility criteria, and corroborated by other two authors; any arising disagreements were solved through discussion among all authors. Risk of bias was negligible, since all publications containing available data were included in the analysis.

### Data collection and standardization

Every data point of interest (i.e. study design, demographics, safety outcomes) was systematically identified and compiled in a specialized, relational SQL database by two authors. For all studies, systemic AE data from the first, second and booster doses were collected. In order to allow a clear readout, only homologous vaccination strategies were included in this booster vaccination assessment; no further filters or limits were applied. Placebo arms were examined where available (i.e. first dose in all three vaccines; second dose for BNT162b2 and mRNA-1273). In all selected studies, solicited AEs (i.e. AEs actively sought after vaccination)^5^ were reported by participants using electronic diaries within seven days of vaccine administration. Solicited systemic AEs were identified and entered into the database. Subsequently, AEs were standardized according to the Medical Dictionary for Regulatory Activities (MedDRA version 22.1, English) and age groups were standardized according to pre-established criteria (Table 3). Only AEs occurring with at least two vaccines were analyzed. The data was standardized in order to identify most comparable subgroups across the three vaccines. The data was analyzed and the results visualized with the software TIBCO Spotfire (version 11.4.0).

## Results

The selection process is depicted in Figure 1. After exclusion of non-approved dosages and dosing schedules, ten publications reporting on clinical trials were identified (Table 1), including seven journal articles and three regulatory (FDA) documents. The entire data compiled in the SQL database amounted to 71 different study arms, corresponding to all information relevant for vaccine safety after first, second and booster doses at the time of analysis.

**Figure 1.**
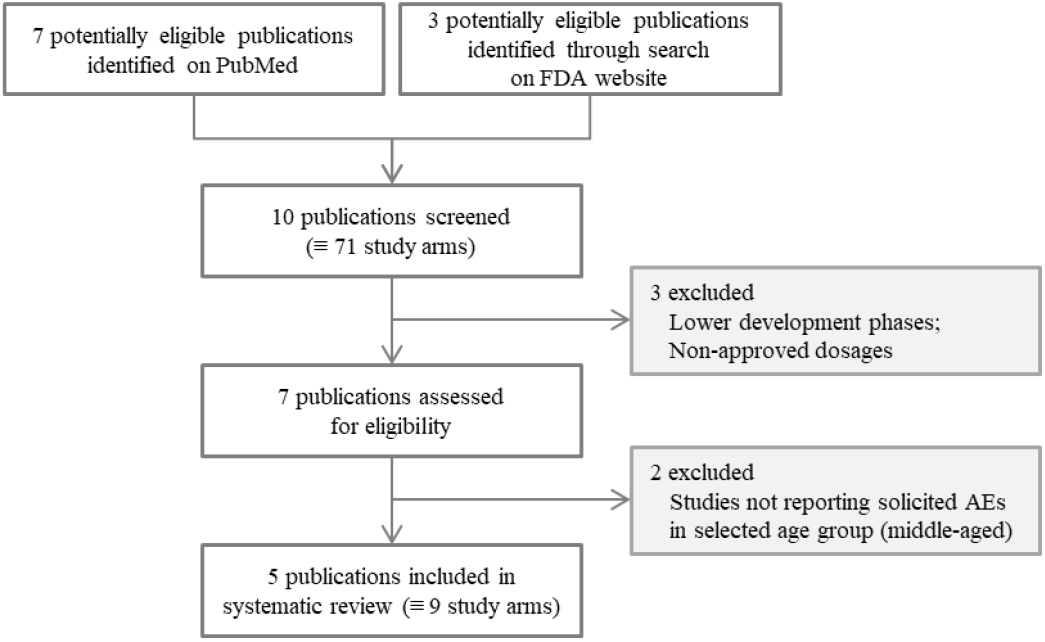
Flowchart showing the selection process for the data to be included in the present vaccine safety outcome assessment (as described in Table 1).

**Table 1.** Stepwise selection of publications (i.e. sources) containing relevant information to be included in the safety assessment of COVID-19 vaccines (first, second and booster dose) approved or authorized in the USA. Ten publications were initially identified and compiled in a SQL database. **A)** All ten identified publications containing relevant information (i.e. prospective data from clinical trials) about the safety of COVID-19 vaccines approved or authorized in the USA (first, second and booster dose). **B)** Standardization step to identify comparable age groups among all trials. **C)** Publications with comparable safety outcomes reported separately for the first, second and booster vaccine doses in middle-aged participants.

| Vaccine | ClinicalTrials.gov ID | Trial phase | (A) Identified data sources | (B) Standardization of age groups | C) Publications with middle-aged subjects |
| --- | --- | --- | --- | --- | --- |
| BNT162b2 | NCT04380701 | Phase I/II | Sahin et al (2020)<sup>6</sup> | ✓ |  |
|  | NCT04368728 | Phase I/II/III | Mulligan et al (2020)<sup>7</sup> | ✓ |  |
|  |  |  | Polack et al (2020)<sup>8</sup> | ✓ | ✓ |
|  |  |  | Walsh et al (2020)<sup>9</sup> | ✓ |  |
|  |  |  | FDA Fact Sheet (Nov 2021) – Biontech<sup>10</sup> | ✓ | ✓ |
| mRNA-1273 | NCT04283461 | Phase I | Anderson et al (2020)<sup>11</sup> | ✓ |  |
|  |  |  | Jackson et al (2020)<sup>12</sup> | ✓ |  |
|  | NCT04470427 | Phase III | Baden et al (2020)<sup>13</sup> | ✓ | ✓ |
|  | NCT04405076 | Phase II | FDA Fact Sheet (Oct 2021) – Moderna<sup>14</sup> | ✓ | ✓ |
| Ad26.COV2.S | NCT04505722 | Phase III | FDA Fact Sheet (Oct 2021) – Janssen<sup>15</sup> | ✓ | ✓ |
|  | NCT04535453 | Phase II |  | ✓ | ✓ |

The data structure showed three trial phases (I–III) (Table 1), various numbers of subjects depending on trial phase (Table 2), and three distinct dose analysis sets (first dose vs. second dose vs. booster dose). Moreover, eleven age ranges were identified across the three analyzed vaccines; a standardization step as shown in Table 3 allowed their clustering into three age groups: Overall, middle-aged, and senior participants. The overall group would best represent the general population; however, data was available in all studies only for the groups of middle-aged and senior participants. Therefore, for this assessment, only the middle-aged group was included, as it is believed to represent the general population better than the senior group (Table 4). Accordingly, five publications were found suitable for safety analysis per administration dose (Table 2), reporting data on five trials: One phase I–III study was assessed for BNT162b2 (reported in one original article and one FDA document), whereas for mRNA-1273 and Ad26.COV2.S one phase II and one phase III study each were included (one original article and one FDA document for mRNA-1273; one FDA document for Ad26.COV2.S). The entire data was compiled in the SQL database, amounting to nine different study arms corresponding to all relevant vaccine safety information for first, second and booster doses at the time of analysis.

**Table 2.** Details of the five publications selected for inclusion in the safety outcome assessment per administration dose after data standardization.

| Label | Dose | Study arm | Number of participants | Age groups | ClinicalTrials.gov ID | Sources |
| --- | --- | --- | --- | --- | --- | --- |
| BNT162b2 | First | Placebo | 10,896 | 16–55 years | NCT04368728 | Pollack et al (2020)<sup>8</sup> |
|  |  | Verum | 10,889 |  |  |  |
|  | Second | Placebo | 10,896 |  |  |  |
|  |  | Verum | 10,889 |  |  |  |
|  | Booster | Verum | 290 | 18–55 years |  | FDA Fact Sheet (Nov 2021) – Biontech<sup>10</sup> |
| mRNA-1273 | First | Placebo | 10,918 | 18–64 years | NCT04470427 | Baden et al (2020)<sup>13</sup> |
|  |  | Verum | 11,406 |  |  |  |
|  | Second | Placebo | 10,918 |  |  |  |
|  |  | Verum | 11,406 |  |  |  |
|  | Booster | Verum | 129 |  | NCT04405076 | FDA Fact Sheet (Oct 2021) – Moderna<sup>14</sup> |
| Ad26.COV2.S | Single-dose | Placebo | 2,049 | 18–59 years | NCT04505722 | FDA Fact Sheet (Oct 2021) – Janssen<sup>15</sup> |
|  |  | Verum | 2,036 |  |  |  |
|  | Booster | Verum | 89 | 18–55 years | NCT04535453 |  |

**Table 3.** Age groups as reported in journal articles and regulatory documents and their standardization for analysis.

| Age as reported [years] | Standardized age groups |
| --- | --- |
| 16–55 | Middle-aged |
| 18–55 |  |
| 18–59 |  |
| 18–64 |  |
| 18–95 | Overall |
| ≥ 18 |  |
| 55–70 | Senior |
| > 55 |  |
| ≥ 60 |  |
| ≥ 65 |  |
| ≥ 70 |  |

**Table 4.** Age groups reported in publications providing safety data for each vaccine. Please note that the overall group would best represent the general population; however, data was available in all studies only for the groups of middle-aged and senior participants. Therefore, for this assessment, only the middle-aged group was included, as it is believed to represent the general population better than the senior group.

|  |  | Vaccines |  |  | Age groups included in assessment |
| --- | --- | --- | --- | --- | --- |
|  |  | BNT162b2 | mRNA-1273 | Ad26.COV2.S |  |
| Age Groups | Middle-aged | ✓ | ✓ | ✓ | ✓ |
|  | Overall |  | ✓ | ✓ |  |
|  | Senior | ✓ | ✓ | ✓ |  |

It must be considered that different numbers of participants were included in the trials for each vaccine (BNT162b2: 21,785; mRNA-1273: 22,324; Ad26.COV2.S: 4,085) (Table 2). In addition, the publications regarding booster vaccinations only reported on rather few participants (BNT162b2: 290; mRNA-1273: 129; Ad26.COV2.S: 89) in comparison to the verum arms of the initial trials. Therefore, different numbers of participants were analyzed for different vaccine doses, representing a statistical limitation of this assessment. The systemic AEs can be divided in three groups according to their frequency with the three vaccines across doses: Particularly common AEs (headache, fatigue), practically non-occurring AEs (fever), and AEs somewhere between the first two groups (arthralgia, myalgia, chills). The most common AEs with the booster dose of any vaccine were fatigue, headache and myalgia, as already seen in the pre-booster doses (Figure 2A). Fever was the systemic AE with the lowest frequency across all vaccines and doses. In the case of the mRNA vaccines (mRNA-1273 and BNT162b2), systemic AEs occurred less frequently after the first dose and more frequently after the second dose. The AE frequencies for the booster dose lied between those of the first and second doses, with the exception of arthralgia, myalgia and fatigue for BNT162b2, which had slightly higher frequencies with the booster. It is noteworthy that the frequency of fever decreased to about half from the preceding dose (i.e. second dose for BNT162b2 and mRNA-1273, first dose for Ad26.COV2.S) to the booster dose for all vaccines. Because the vector vaccine Ad26.COV2.S only requires a single initial dose, we compared the data for the first dose with that of the booster vaccination. With the exception of fatigue, the systemic AE rates were lower with the booster dose than with the first vaccine dose. No data is available for Ad26.COV2.S on chills and arthralgia (Figure 2A).

**Figure 2.**
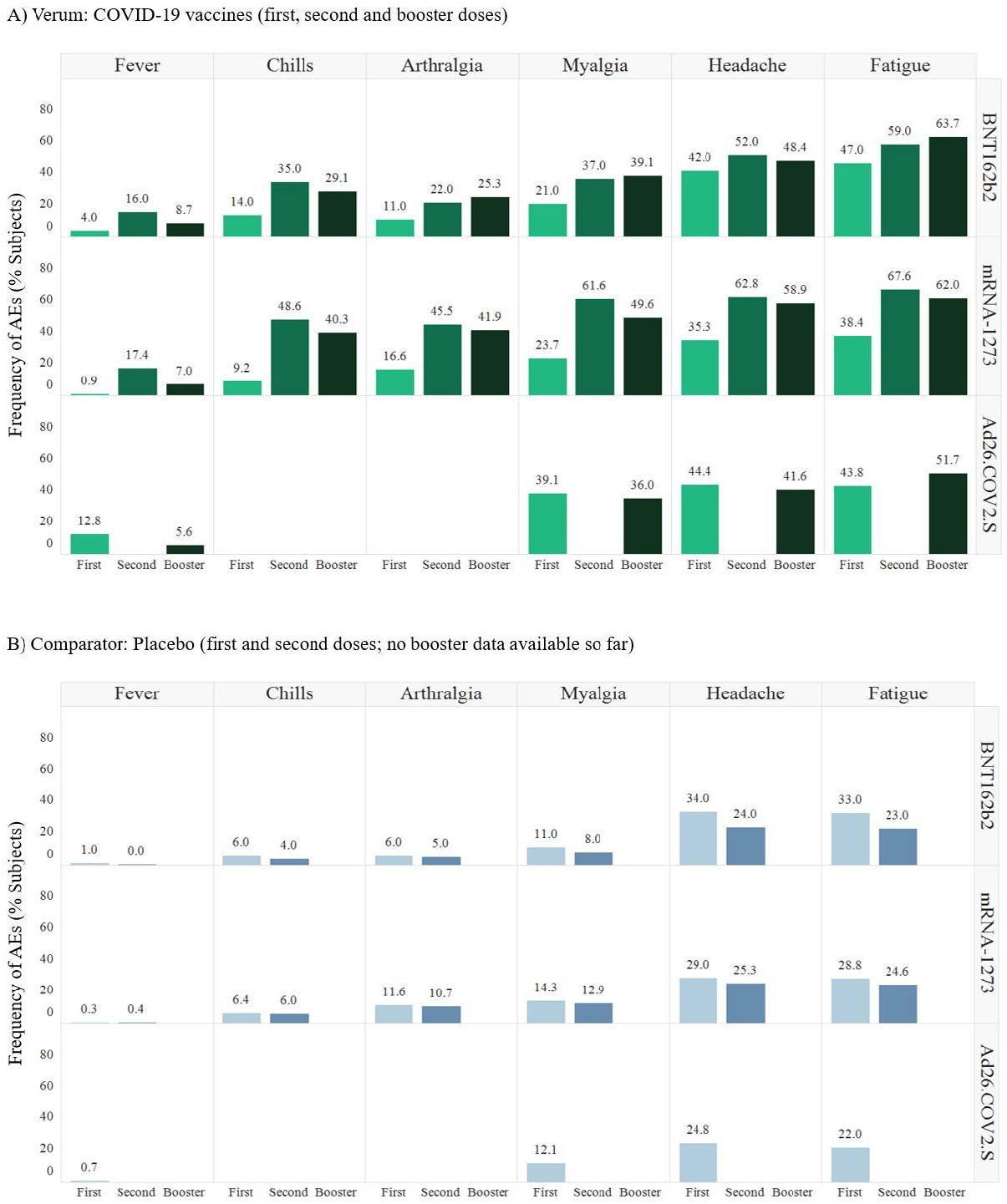
Most common systemic adverse events occurring after the first, second and booster doses of anti-SARS-CoV-2 vaccines in **A)** verum arms and **B)** placebo arms. The highest frequencies for each adverse event occurring with each vaccine are shown. No data was reported for chills or arthralgia for Ad26.COV2.S. The Janssen vaccine is approved with a single dose before the booster dose.

To further illustrate the differences between the individual vaccine doses, we plotted the changes in the frequency of AEs for all vaccines. The comparison of the frequencies between the first and second doses showed that the AEs for the mRNA vaccines increased by 10% to about 39% (Figure 3A), but decreased for almost all AEs in the placebo arms between the same doses (Figure 3B). When comparing the frequencies of the AEs associated with the booster dose with those of the previous doses (i.e. second dose for BNT162b2 and mRNA-1273, first dose for Ad26.COV2.S) (Figure 3C), all rates decreased for mRNA-1273 (clearly with myalgia by −12%). BNT162b2 displayed similar results for the booster and the second dose (slightly lower for chills and headache; slightly higher for arthralgia, fatigue and myalgia). For Ad26.COV2.S, a slight increase was noted in the frequency of fatigue from the previous dose to the booster dose (7·9%).

**Figure 3.**
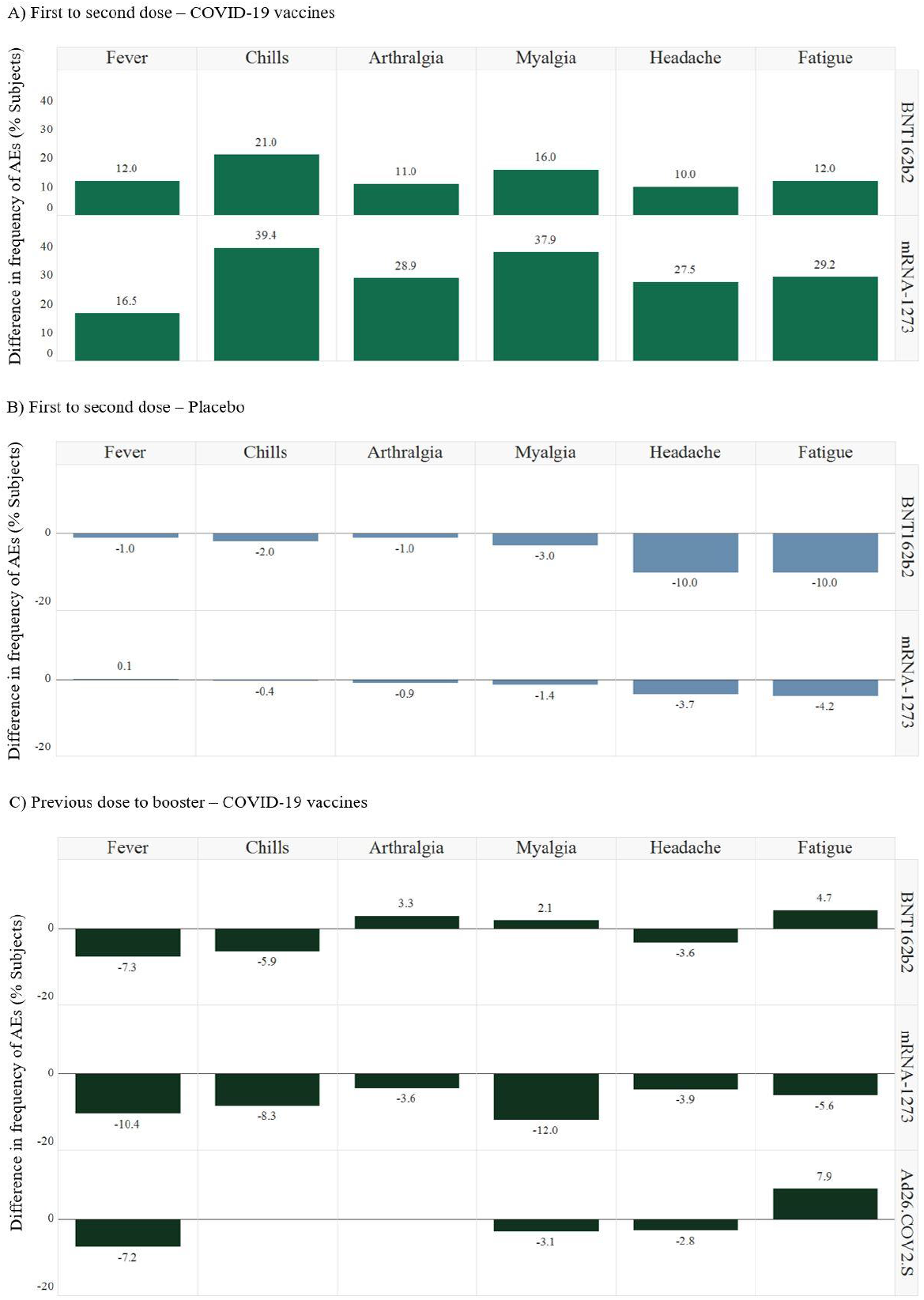
Differences in the frequencies of most common systemic AEs between doses: **A)** from first to second vaccine dose in the verum group, **B)** from first to second vaccine dose in the placebo group, and **C)** between booster and previous dose (i.e. second dose for BNT162b2 and mRNA-1273, first dose for Ad26.COV2.S). The highest frequencies for each AE occurring with each vaccine are shown. No data was reported for chills or arthralgia for Ad26.COV2.S. The Janssen vaccine is approved with a single dose before the booster dose.

High frequencies were observed for most of the systemic AEs in the placebo arms, with the exception of fever, which was practically absent in all three vaccines (Figure 2B).

If the frequencies of the individual systemic AEs in the placebo arms are subtracted from those in the vaccine arms, the ranges within each vaccine remain similar except for fever (first dose: 5–14% for BNT162b2, 2·8–9·6% for mRNA-1273, 19·6–27% for Ad26.COV2.S – Figure 4A; second dose: 17–36% for BNT162b2, 34·8–48·7% for mRNA-1273; no second dose for Ad26.COV2.S – Figure 4B). With this approach, the main AE frequency groups previously described are no longer detectable.

**Figure 4.**
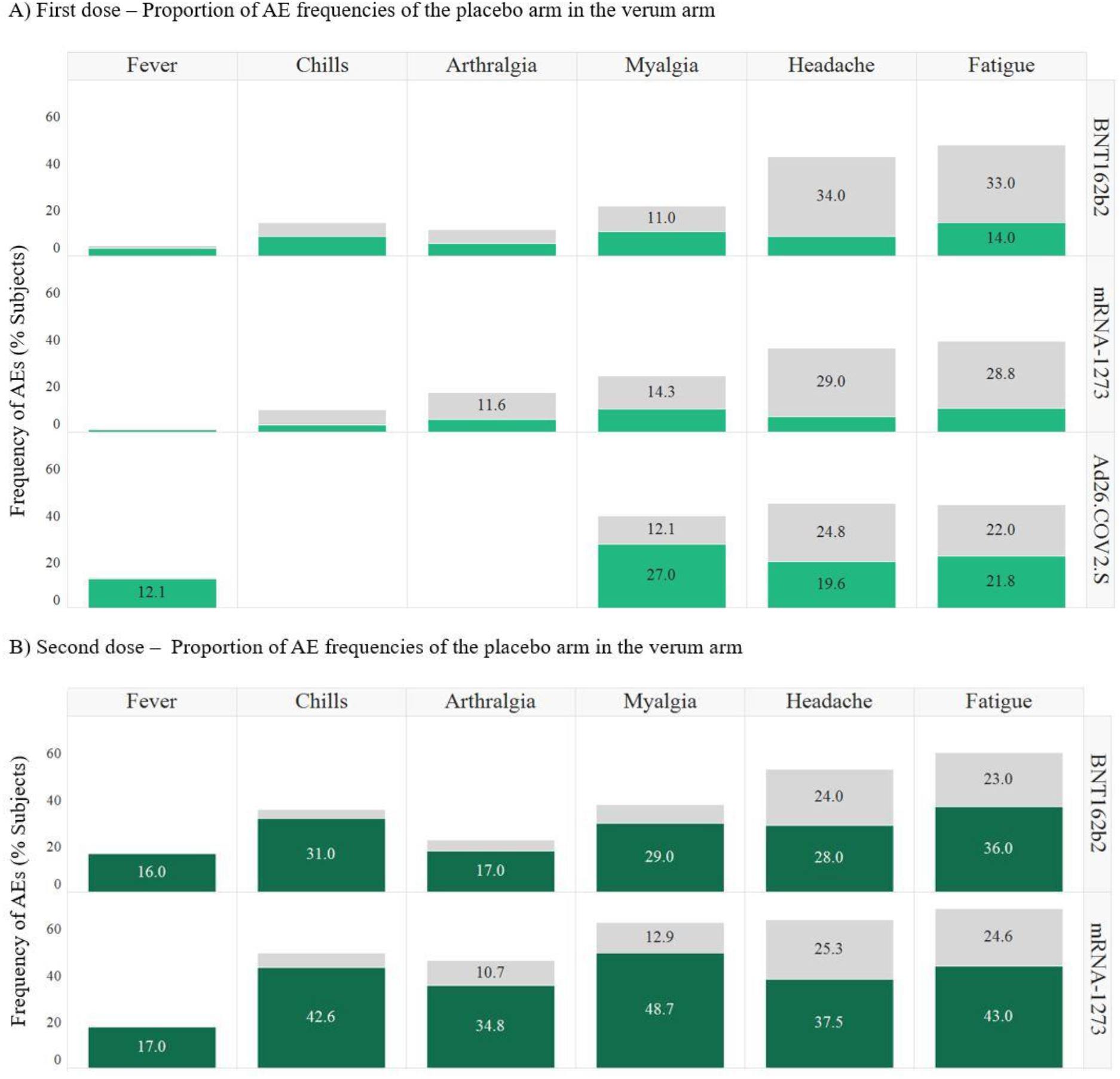
Graphic visualization of the nocebo effect in COVID-19 vaccination. The values for AE frequencies in the placebo arm in relation to the verum arm are represented in **A)** with the first vaccine dose and in **B)** with the second vaccine dose; Ad26.COV2.S is approved without a second dose. The gray portion represents the AE percentages in the placebo arm; the green portion corresponds to the remaining proportion of AEs occurring in the verum arms after subtraction of the values observed for the same AEs in the placebo arm.

## Discussion

Comparisons between different clinical trials can have a major impact on both individual treatment decisions as well as on decisions during the pharma R&D process. However, head-to-head comparisons are rarely conducted; instead, indirect comparisons are performed (e.g. Cochrane analyses, review articles). Here, we indirectly compare trial data for COVID-19 vaccines which currently have an EUA in the USA for booster vaccination. We investigated all systemic, solicited AEs (i.e. those AEs actively sought after vaccination)^5^ reported for each vaccine after the first, second and booster dose.

Overall, the systemic AEs can be divided in three groups according to their frequency with the three analyzed vaccines across doses: Particularly common AEs (headache, fatigue), practically non-occurring AEs (fever), and AEs with a frequency somewhere between the first two groups (arthralgia, myalgia, chills). Upon subtracting AE frequencies observed with placebo from those reported with the vaccines, these main AE frequency groups are no longer detectable.

Safety outcomes after the first and second vaccine doses have been reported recently^3^. Overall, as for the first and second doses, the authorized booster vaccine doses are well tolerated, and most AEs are only mild. Fatigue, headache and myalgia prevailed as the most common AEs. Fever was the AE with the lowest frequency across all vaccines and doses.

Various patterns could be verified in the AE frequencies. In the case of the mRNA vaccines (mRNA-1273 and BNT162b2), systemic AEs occur less frequently after the first dose and more frequently after the second dose. With few exceptions, the frequencies for the booster dose lie between those of the first and second doses. For Ad26.COV2.S, only one initial dose is needed. Most systemic AEs are lower with the booster dose than with the first dose.

AEs arising from drug therapy are the most common reasons provided by patients who do not accept medication or fail to adhere to treatment. Whereas this may primarily have individual consequences, in the case of a pandemic the refusal to accept vaccination rises to a social problem^16^.

For the analyzed COVID-19 vaccines, the nocebo effect with mRNA-1273 and BNT162b2 decreased from the first to the second dose for all subject-reported systemic AEs; there were almost no fever cases in any of the three placebo arms. One can speculate whether the nocebo effect would decrease even more significantly with a booster-placebo control arm.

In contrast to all other common systemic AEs, fever was barely reported in the placebo arms. A possible explanation for this is that fever can be measured objectively. In contrast, all other assessed systemic AEs are subject-reported, and therefore prone to a negative perception bias. This observation is in accordance with a nocebo effect, defined as AEs occurring during sham treatment. The nocebo effect has been reported in usual drug treatments, particularly for pain^17^. In addition, it has been recently reported for several AEs in COVID-19 vaccination^2^. The nocebo effect in vaccination is of particular interest since a high frequency of non-objectively measurable AEs may negatively impact vaccination willingness^18^. Therefore, fever as an objectively measurable parameter could help to better understand the extent of the nocebo effect of subjective AEs during vaccination. It is interesting that the frequency of fever decreases to about half with the booster dose of the three vaccines in comparison to the respective previous dose. In contrast, the frequencies of the other systemic AEs only decrease slightly in the same comparison, and for some AEs these values even increase. Even though distinct AEs may develop differently between vaccine doses, it can be speculated that the frequency of all systemic AEs, also with the booster dose, may be strongly influenced by a nocebo effect. Ultimately, this question can only be answered with placebo-controlled vaccine studies of the booster dose.

The systemic review presented here is only a small excerpt from a larger systematic COVID-19 vaccine safety assessment based on the most relevant trial data (safety and efficacy). The multidimensional assessment of vaccine data presented here may serve as basis for a public awareness campaign to combat hesitancy in receiving a booster dose of the approved anti-SARS-CoV-2 vaccines.

## Data Availability

All data produced in the present work are contained in the manuscript.

## Limitations of the study

The trials analyzed here included considerably different numbers of participants, in particular for the booster dose, representing a statistical limitation of this assessment.

## Contributors

CB and JL designed the study. AG and JL searched and identified the relevant data sources. AG, JM and TG selected the articles and extracted the data. AG, CB and MSM validated the compiled data. CB, JL and MSM analyzed the data and prepared data visualization. JL and TG wrote the original draft of the manuscript; AG, CB, JM and MSM critically revised the final version. CB, JL and TG supervised the entire project.

## Declaration of Competing Interest

The authors declare that the analysis was conducted in the absence of any commercial or financial relationships that could be construed as a potential conflict of interest.

## Data availability statement

The present systematic review was not registered in any international database.

## Notes

### Competing Interest Statement

The authors have declared no competing interest.

### Funding Statement

This study did not receive any funding.

